# Findings from a Pilot of a Near Real Time Disease Surveillance Tool on the NATO Mission in Kosovo

**DOI:** 10.1101/2023.08.21.23293305

**Authors:** Robert Lindfield, Claudia Biendl, Mate Toth, Silke Ruhl

## Abstract

NATO has a requirement for a near real time surveillance (NRTS) tool to rapidly identify disease outbreaks but no deployable tool is available. The NATO Centre of Excellence for Military Medicine (MILMED COE) developed a NRTS tool and piloted it on the NATO Mission in Kosovo (KFOR).

Ten medical treatment facilities (MTFs) in KFOR piloted the tool which is an app on a smart device. All first presentations were eligible for inclusion into the app. Primary presenting symptoms were mapped to one or more of 40 symptoms. No patient identifiable information was collected. Analysis was available to each MTF and the Medical HQ staff (JMED) which provided a summary of the data collected. Combinations of symptoms resulted in an alert email to the JMED and MTF.

An evaluation was conducted at two time points consisting of data analysis and semi-structured interviews with key informants from each participating MTF and JMED team.

1351 patients were entered during the pilot. 851 reports had symptoms recorded. The leading symptoms were pain-in-throat, cough and temperature. Two investigations were launched based on reporting in the app; a cluster of skin lesions and two cases of diarrhoea. No significant outbreaks were reported during the pilot or detected by the app.

Interviews revealed that the app was easy to use and the symptom list was comprehensive. The analysis tool was not regularly viewed by MTFs however the JMED team used it frequently. Alerts were considered extremely valuable by both the MTFs and JMED team.

The KFOR pilot provided an excellent opportunity to pilot the tool in a NATO Mission and established that the way it collected data and alerted issues worked well. Further validation and analysis of the tool is required before it can be rolled out further.

## Introduction

NATO first described the requirement for a disease surveillance function at the Prague Summit in 2002 [1]. This was a response to the deliberate release of anthrax in the US during 2000 and aimed to enhance the Alliance’s defence capabilities against weapons of mass destruction. Since 2002 there have been several attempts by Nations to introduce near real time disease surveillance tools including by the French [2] and the British [3] however these are no longer in use.

The only disease surveillance tool currently deployed on NATO missions is EpiNATO-2 which collects weekly syndromic data from medical treatment facilities in Missions [4]. The tool reports broad syndromic categories based on the patient’s main presenting condition. Evidence suggested that EpiNATO-2 was unable to detect outbreaks or incidents of infectious disease or deliberate release due to its weekly reporting interval which drove the development of the NRTS Tool to address this requirement [5]. This was aided by improvements in software required to deliver this type of capability.

A tool was developed using the Microsoft 365 suite of products. This primarily focused on using MS PowerApps and PowerBI to collect, collate and display results and generate alerts, and Sharepoint to share and store data. The tool consisted of three elements; a data entry app, a method of sharing data including alerts, and a way of collating and displaying the information.

The symptom list was agreed by an expert group from the NATO Force Health Protection Working Group. A consensus process using a French surveillance tool was developed and a list of symptoms and signs thought appropriate to identify an incident or outbreak of infectious disease or deliberate release was approved.

The tool was initially tested on the NATO Coalition Warrior Interoperability Exercises in 2021 and 2022 which established that the tool was able to share data with National and NATO systems. The tool underwent clinical piloting during the 2022 NATO CLEAN CARE Exercise and, following further development, was considered ready for a live pilot.

This paper describes the results of the live pilot of the Near Real Time Surveillance Tool in the NATO Mission in Kosovo.

## Methods

### Approval

Permission was granted for the pilot from the Commander of the NATO Mission in Kosovo (KFOR). No patient identifiable information was collected. This was considered as a service evaluation as it ran alongside EpiNATO-2 data reporting on KFOR. All MTF specific data has been anonymised in this report. For further information please contact the lead author.

### Technical Elements

Data was entered into the NRTS Tool using a bespoke app on a handheld device such as a smartphone or tablet. For the KFOR Pilot every MTF was provided with a tablet to collect data.

All clinical teams received a standardised training package at the beginning of the pilot with a refresh or training for new MTFs at the midpoint of the pilot. The package consisted of an explanation and demonstration of the app and the analysis page. This was complemented by a list of dummy patients (see Supplementary Information) which each team could use to practise entering data. Each team was provided with a tablet and a set of documentation including the standard operating procedure (see Supplementary Information), the dummy patients and contact details should there be any questions or issues.

The MTF was requested to enter the primary presenting symptoms of any patient attending. The clinician could choose one or more of forty different symptoms and signs on the app. For patients with symptoms not on the app the clinician was requested to tick a ‘symptom not included’ box and enter text to describe the main presentation. Non-first presentations were also recorded.

When the tablet was connected to the internet then the information was automatically uploaded to Sharepoint. When the tablet was not connected then the information was held on the device until a connection was available and then uploaded in a single burst. The list of symptoms and signs is included in Supplementary Information.

The data was used to populate the PowerBI analysis page which displayed a range of information including the number of patients and types of presentation by MTF, Camp and Mission. Each MTF was permitted to view their own data and the data for the camp in which they were located. The Joint Medical Team (JMED) at the Mission Headquarters was able to see all data.

Alerts could be generated by the software. An alert email was sent to pre-determined email addresses based on the presence or absence of certain conditions including the number of cases, types of symptoms or location of the MTF.

All results were made available to every MTF and the JMED on an analysis page within a separate app on the tablet. The analysis page showed the number of patients entered into the app, the top five symptoms reported and provided a detailed list of the symptoms reported for every patient seen by each MTF throughout the pilot period.

### Setting

During the pilot there were five camps in KFOR dispersed over a wide geographical area. For the purpose of the pilot the three camps with the largest populations and most MTFs were chosen. This was expanded to include an additional camp for the second phase of the pilot at the request of the JMED team. Table 1 shows the amount of time each MTF contributed to the pilot.

**Table 1:**
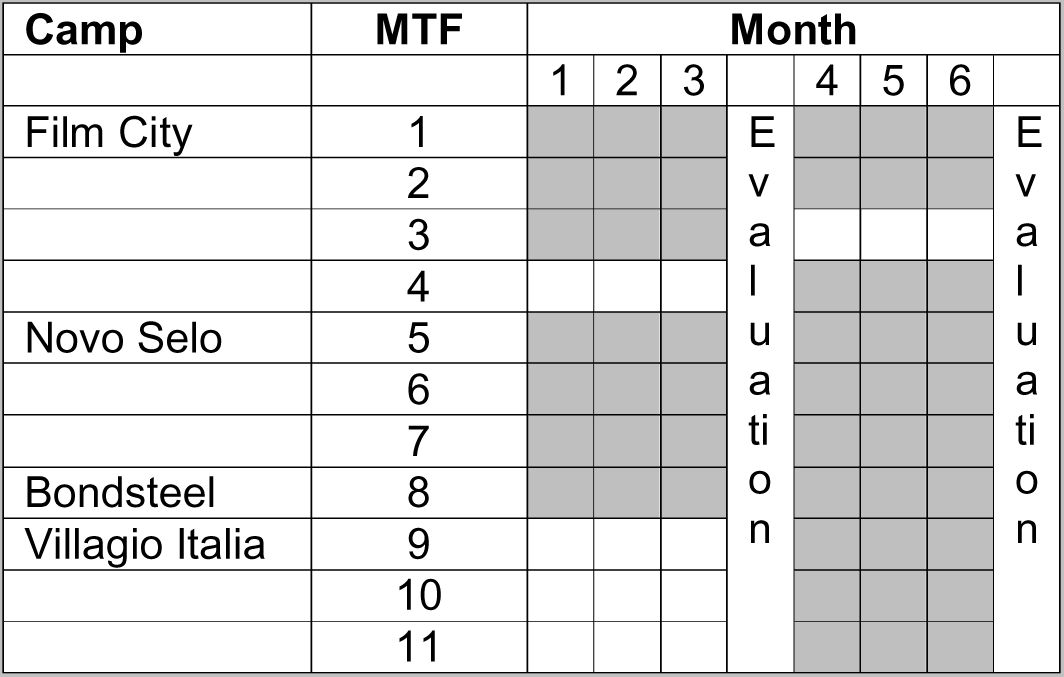
Involvement in the Pilot by MTF.

### Evaluation

There were two evaluation time points. The pilot was originally planned to finish after three months so an evaluation was arranged for that time. However, there was agreement that the pilot could continue for a further three months so additional MTFs could be recruited and different elements of the tool tested. The second evaluation time point was at the end of six months.

The evaluation consisted of face-to-face interviews with key informants; the individuals responsible for data entry in each MTF, their colleagues in the MTF as required, and the JMED Team.

An a-priori question set (see Supplementary Information) was developed addressing three main elements; the utility of the app, the analysis page and the future use of the tool. These were framed as a series of open-ended questions and shared with all participants two weeks in advance of the interviews so that they could be discussed and translated if required.

The interviews were arranged through the JMED Team and timed based on clinical commitments. Attempts were made to reach key informants by phone if they were not available at the agreed time.

The evaluation was supported by data on the use of the tool including the number of patients entered by each MTF and the leading symptoms. It was also possible to count the number of times each MTF had accessed their own analysis page using the PowerBI system.

### Completeness of data entry

It was not possible to review patient records in each MTF however all MTFs also collected EpiNATO-2 data throughout the pilot. Whilst not being directly comparable due to the data collection categories, noting that EpiNATO-2 is syndromic not symptom-based, it was possible to compare the number of entries for specific categories such as gastrointestinal infection (EpiNATO-2) and comparing this with the number of patients reported in the abdominal symptoms and signs section of the NRTS tool (including vomiting, nausea, diarrhoea, watery diarrhoea, abdominal pain and blood in stool).

### Alerts

An alert was generated to inform the JMED and/or MTF when certain criteria had been met by the data entry that warranted further investigation. The approach during the first part of the pilot was to combine 2-3 symptoms into an algorithm that defined a specific condition – for example; a patient with cough, fever and myalgia would be defined as having influenza-like illness and an alert would be generated to report this. However, when this was trialled in the NATO CLEAN CARE Exercise there were significant concerns about the sensitivity (true positive rate) of this approach as a patient needed to report all three symptoms to generate the alert thereby making it more likely that it would miss people who reported only one or two symptoms.

A solution was developed where the symptom reported became less important than the number, timing and location of patients presenting with the symptom. This alert was tested during the second part of the pilot in KFOR and was generated when two or more patients presented to the same MTF with the same symptom within the same 24 hours. There was also an ‘extension’ alert which was generated when at least one further patient presented with the same symptom in the same camp.

### Analysis

Simple descriptive counts were used to assess the presentations to each MTF. This included new vs follow up, day of presentation, primary presenting symptoms and combinations of symptoms. Additionally, comparison was made between the number of EpiNATO-2 reports and the number of NRTS reports.

The interviews were transcribed by hand to capture key elements of the feedback. The interviews took place in English which was, frequently, not the first language of the interviewee and, as a result, the notes were not exhaustive. Simple thematic analysis was used to evaluate the interviews. It should be noted that, as this was a service evaluation, a formal mechanistic process for analysing qualitative data was not appropriate however themes were developed from the notes and discussion amongst the lead evaluators (RL and CB).

## Results

1697 patient presentations were entered into the software during the pilot. Of these, 1351 (79.6%) were classified as first presentations. Chart 1 details the total number of reports, both first and subsequent presentations, by day. Four peaks in reporting were seen; 17 Nov, 07 Dec, 20 Jan and 14 Feb (see Chart 1). The number of first presentations reported varied through the pilot with most non-first presentations in the first three months (see Chart 1). The majority of non-first presentations (255/346 (74%)) were from a single MTF (KFOR_08).

**Chart 1:**
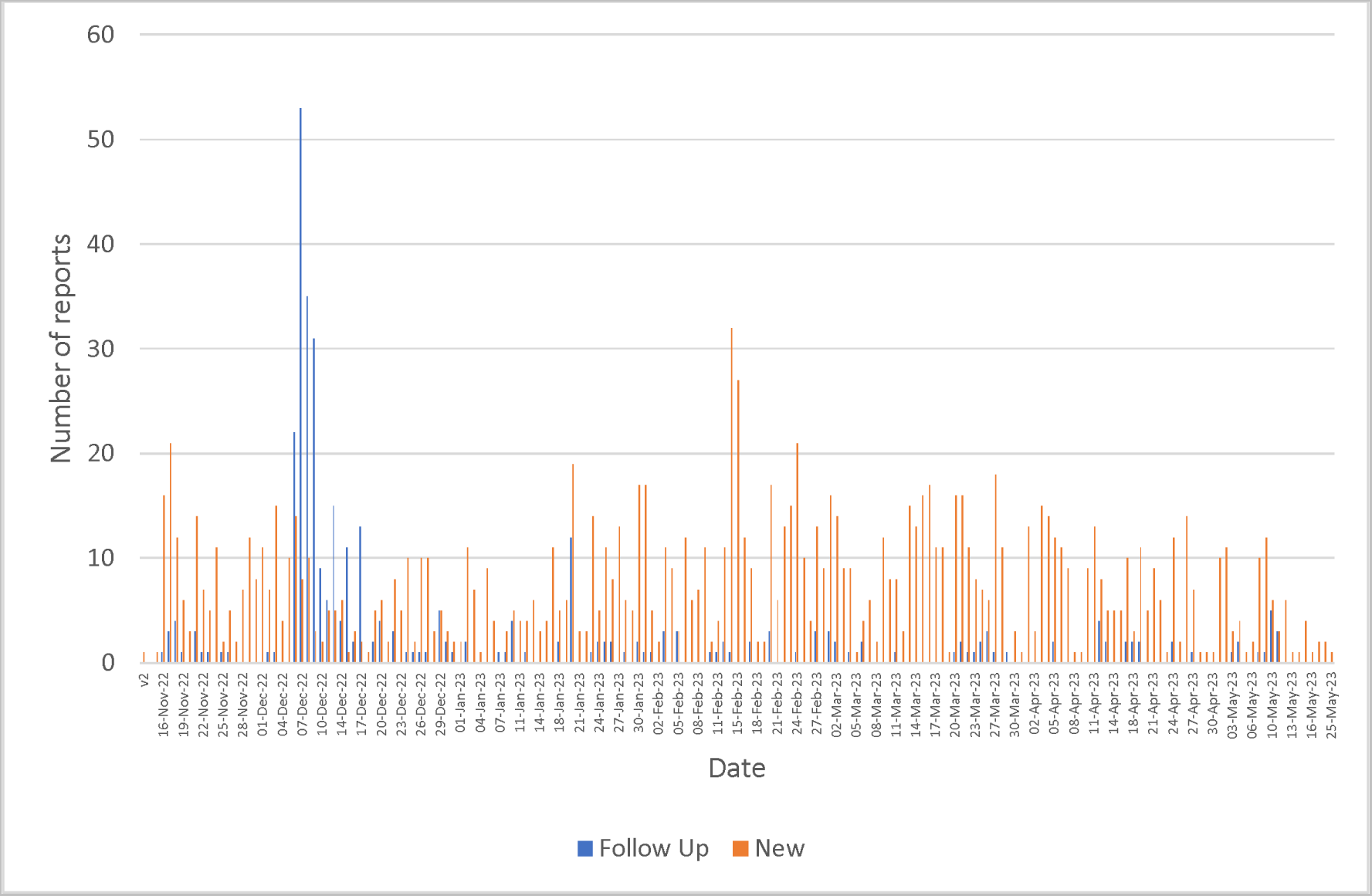
Total number of presentations by date (new and follow up)

It should be noted that the peak in follow-up presentations seen on 07 Dec 22 was reported as Vaccination (n=52). The highest number of new patients reported in one day (n=32) was on 14 Feb 23 which coincided with the first evaluation visit.

### Symptoms

There were 851 reports with symptoms recorded in the app which was 50.2% of all records. Chart 2 lists the frequency of the main symptoms recorded. Pain in throat and cough were the leading symptoms recorded with over twice the number of records compared to the next most frequent symptom which was temperature. It was also possible to determine the frequency and combination of multiple symptoms recorded (see Table 2). The most common combination of symptoms was cough and pain in throat (84 reports) followed by cough and headache (11 reports).

**Chart 2:**
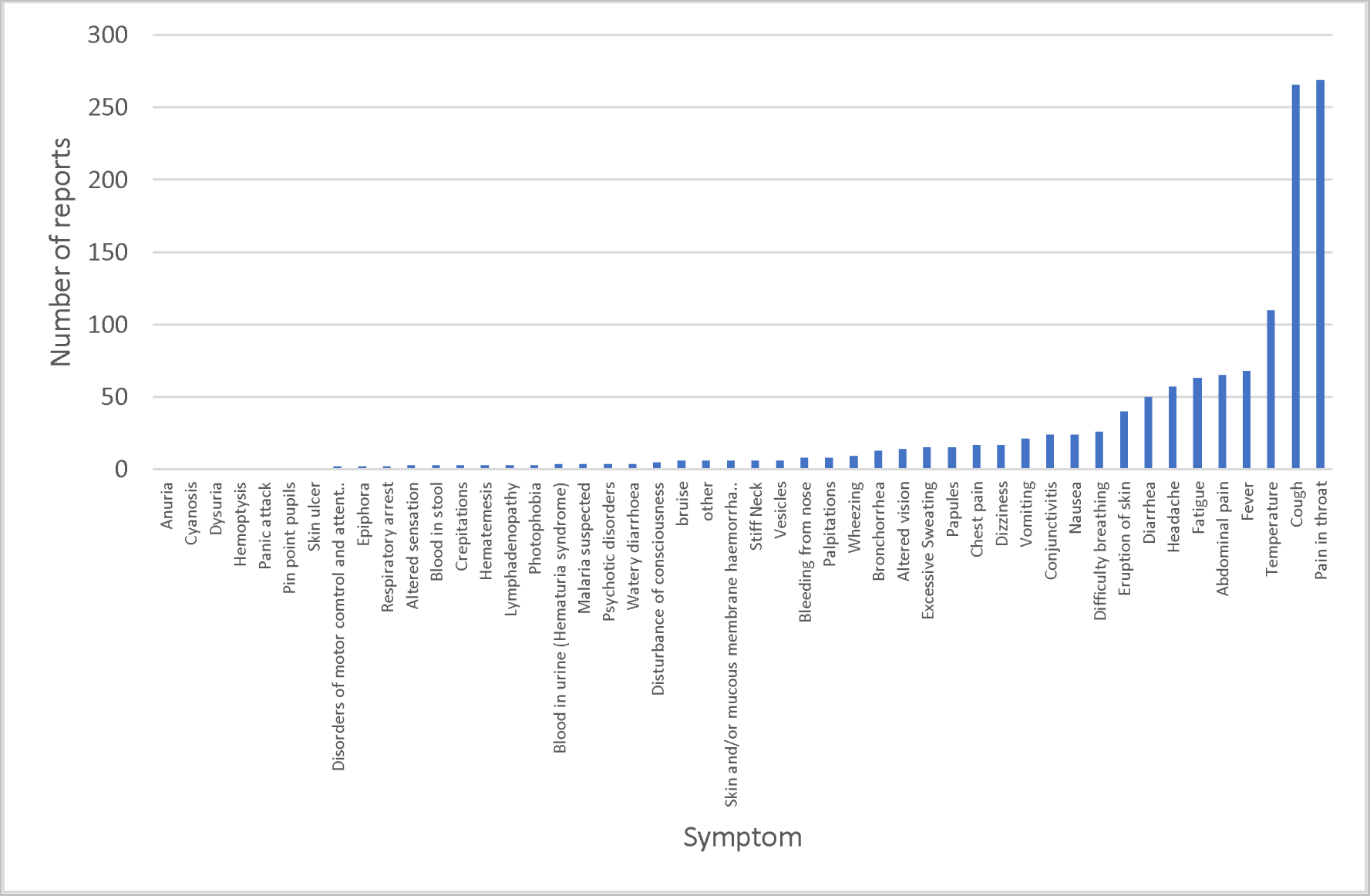
The number of reports of different symptoms.

**Table 2:**
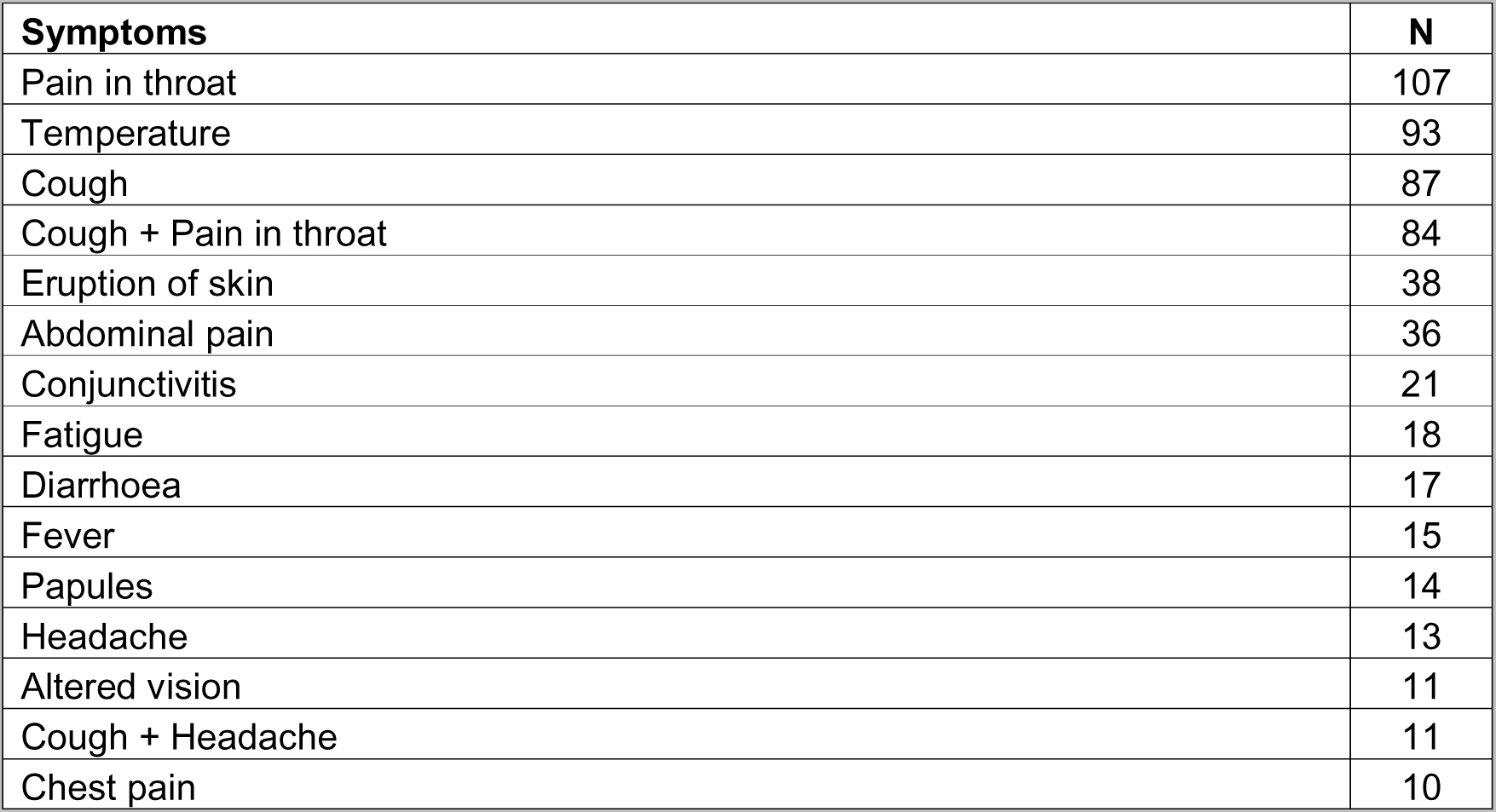
List of most frequent symptoms and combinations of symptoms.

| **Symptoms** | **N** |
| --- | --- |
| Pain in throat | 107 |
| Temperature | 93 |
| Cough | 87 |
| Cough + Pain in throat | 84 |
| Eruption of skin | 38 |
| Abdominal pain | 36 |
| Conjunctivitis | 21 |
| Fatigue | 18 |
| Diarrhoea | 17 |
| Fever | 15 |
| Papules | 14 |
| Headache | 13 |
| Altered vision | 11 |
| Cough + Headache | 11 |
| Chest pain | 10 |

### Days of Reporting

Chart 4 shows the number of reports per day each week. Monday was the most frequent day to report and reporting tailed off through the week with Sunday having the lowest number of reports.

**Chart 4:**
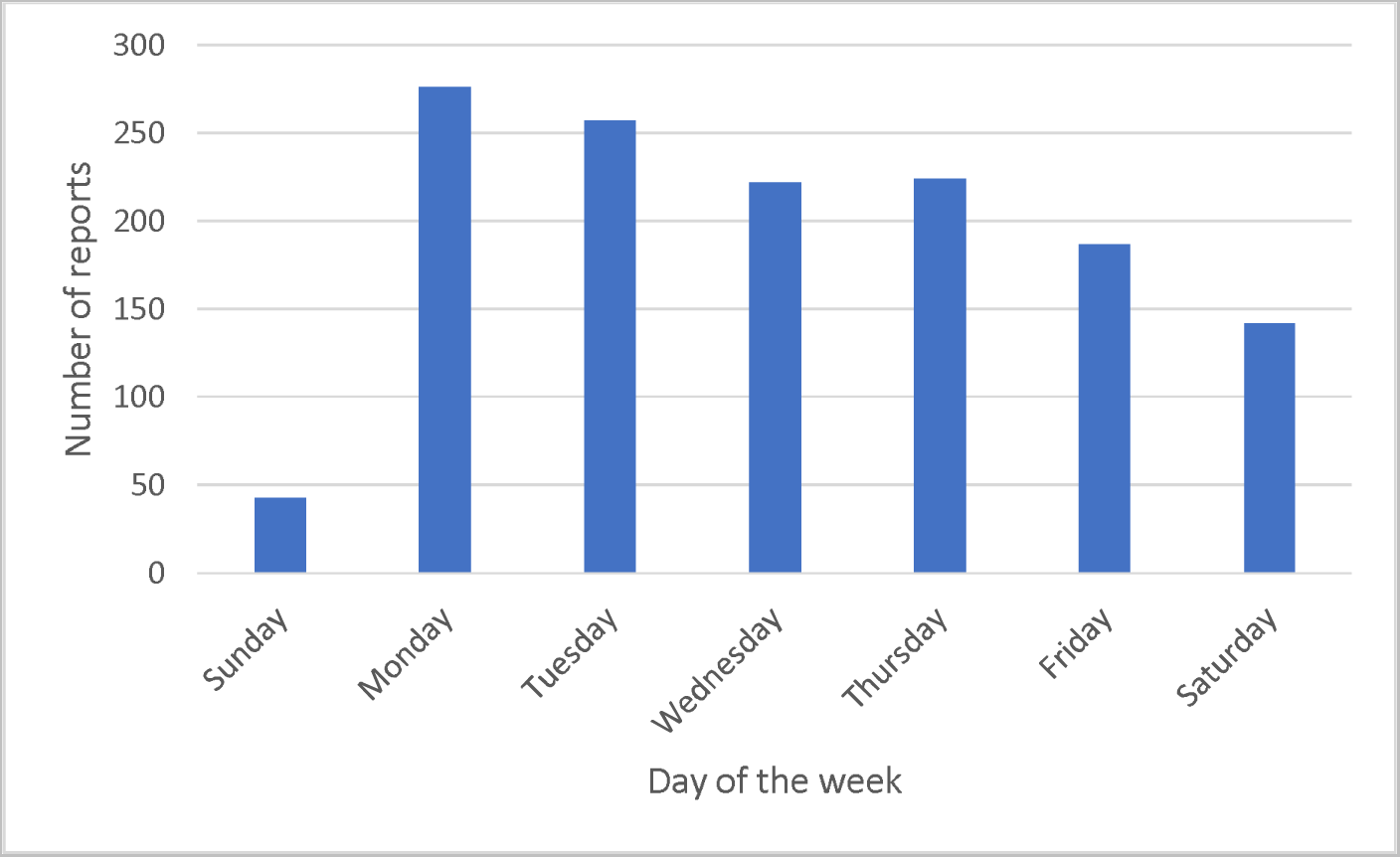
Number of reports by day of the week.

### Reports by MTF

There was variation in the total number of reports received from each MTF (see Chart 5). KFOR_04 and KFOR_12 provided the highest number of reports which was expected as they had the largest population at risk (PAR). It should be noted that it is not possible to report the PAR or any denominator data as this is restricted. The MTFs that were recruited in the second part of the pilot are coloured in red. As expected, they reported fewer patients.

**Chart 5:**
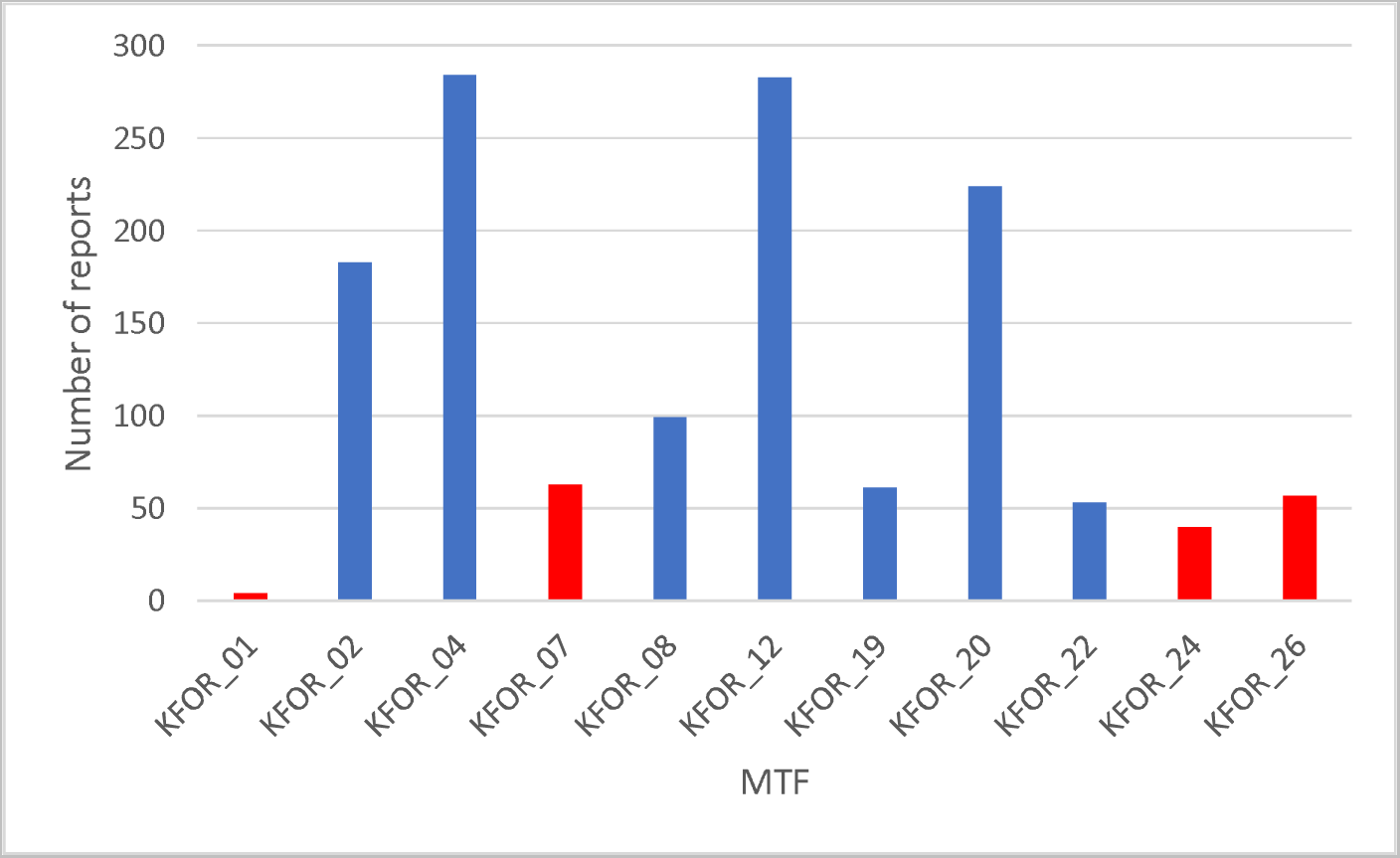
Number of reports by Medical Treatment Facility.

### Evaluation

None of the clinical staff who were trained to use the tool at the start of the pilot were present at the end of the pilot. This meant that there could be an additional assessment of the handover of the app to a new team as part of the evaluation.

### Use of the App

Without exception the app was reported as being easy to use. This was evidenced by the ongoing use of the app when clinical teams in the MTF changed over with no formal handover or training from the MILMED COE team. The app was described as self-explanatory and took only a short time to enter data. Two issues with the app were reported. Firstly, it was only available in English so at least two MTFs translated the symptom list to make it easier to understand. Secondly, the MTFs were still expected to collect EpiNATO-2 data which involved a manual data extraction and there was a request to collect EpiNATO-2 data using the tablet. No-one had any suggestions on how to make the app easier to use.

Clinical teams were asked whether there should be any additions to the symptom list on the app. They consistently asked for musculoskeletal injury be added as this was the largest patient group they reported seeing. Collecting injury data was not the purpose of the tool and this request suggested that the purpose of the tool was not clear to clinical teams. This was the case when medical teams did the handover and not the MILMED COE team.

There were no other symptoms that the clinical teams recommended should be included in the tool. There were also no symptoms that the clinical teams suggested should be removed from the list.

No-one reported discussing the analysis page within their team. It was unclear why this had not happened although several clinicians reported that there was nothing of concern so there was ‘no need to look at the analysis’ however some also reported that they were unaware that the analysis app existed – particularly those who had the tool handed over to them without training. Those that did report looking at the analysis app said that they struggled to understand the analysis page. The analysis page was changed following the mid-term evaluation in an attempt to make it easier to understand but the feedback during the final evaluation was very similar suggesting that the analysis page did not display what the clinical teams wanted to see or that they were not interested in seeing it with several individuals commenting that their role was to enter the data.

The JMED Team had a different perspective from the clinical teams. They were based in the HQ and had responsibility for oversight of the whole Mission. The analysis app provided them with information that they did not have previously and they reported that it facilitated discussion with the MTFs that they would not have had without it. For example, there were several cases of skin rash reported from an MTF that had not previously engaged with the JMED Team. This allowed contact to be established and a discussion on the reasons for the presentations with skin rashes.

The JMED Team provided extensive feedback about the Analysis page during the final evaluation and recommended extensive changes to make it useful. These included minimising the quantity of information being displayed but having access to more detail if required.

### Alerts

Alerts were introduced during the second three months of the pilot in KFOR. There were some technical issues in their development meaning that there were only a few weeks when they were being used. However, their utility was welcomed with feedback by the JMED Team stating their importance.

### Investigations

The JMED Team led two investigations as a direct result of the NRTS reporting. The first investigation involved three reports of ‘papules’ from KFOR_08 on 27 Nov 22. However further investigation revealed seven patients with the primary presenting symptom of ‘papule’ between 17 Nov and 28 Nov 22 (see Chart 5). This led to a further investigation about potential exposures that might have resulted in a skin lesion.

**Chart 5a:**
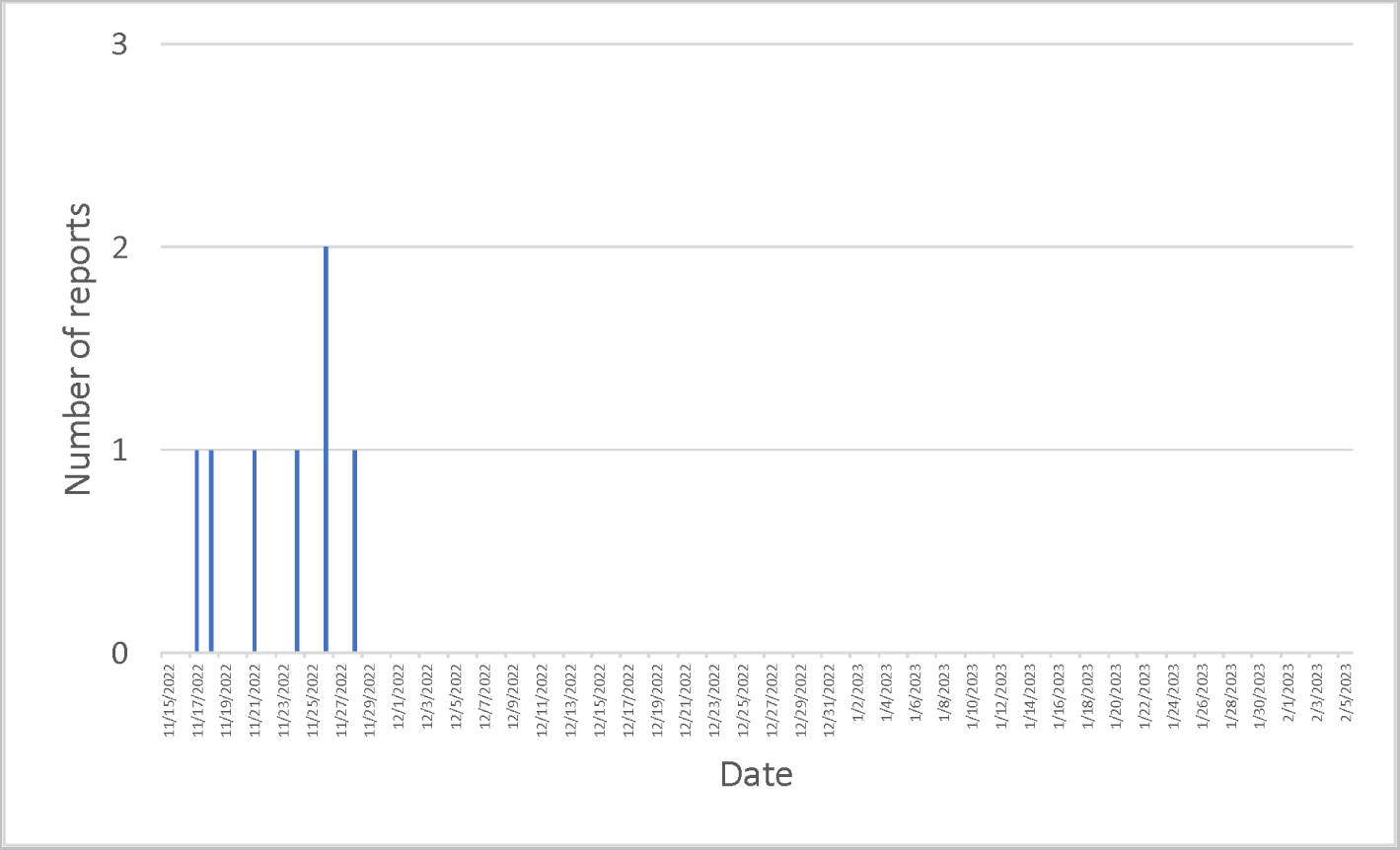
Number of patients reporting with the symptom ‘papules’ to KFOR_08.

The second investigation was triggered by an alert for three patients presenting to KFOR_19 with diarrhoea. Chart 6 shows the number of cases of diarrhoea reported by KFOR_19 for the duration of the pilot. It can be seen that there was a small cluster of cases around 24 Apr 23 which were investigated and found to be associated with a food provider outside the camp.

**Chart 6:**
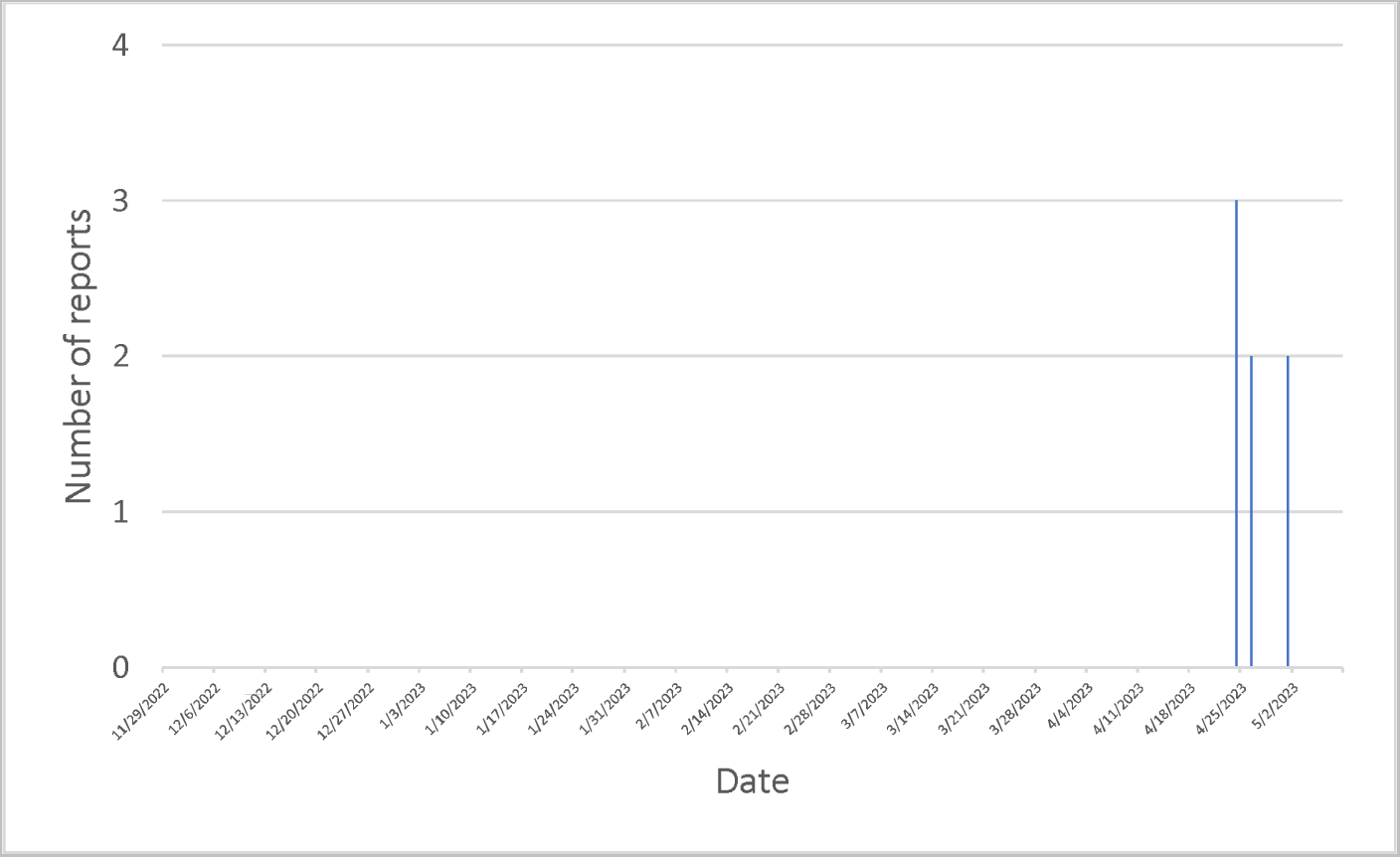
Number of patients reporting with the symptom ‘diarrhoea’ to KFOR_08.

### Validation

Limited ways of validating data entry into the NRTS app were available due to patient confidentiality and lack of access to the MTF’s patient record. However, it was possible to compare the EpiNATO-2 data entries by MTF with the data entered into the NRTS app across the time period of the pilot. This is only indicative as the categories in EpiNATO-2 are broad syndromes (e.g. gastrointestinal infection) which means they cannot be compared directly to NRTS symptoms. To overcome this the symptoms that were most likely to be categorised as gastrointestinal by NRTS (diarrhoea, abdominal pain, vomiting) were grouped and compared to ‘gastrointestinal infection’ - see Table 3 which shows a significant different with the majority of MTFs having fewer entries in NRTS. This suggests underreporting in NRTS but there should be significant caution when interpreting these results.

**Table 3:** Comparison of Gastrointestinal Infection entries between EpiNATO-2 and NRTS.

|  | EpiNATO-2 | NRTS | % |
| --- | --- | --- | --- |
| KFOR_02 | 43 | 12 | 28% |
| KFOR_04 | 33 | 16 | 48% |
| KFOR_08 | 19 | 6 | 32% |
| KFOR_12 | 61 | 11 | 18% |
| KFOR_19 | 13 | 9 | 69% |
| KFOR_20 | 33 | 17 | 52% |
| KFOR_22 | 1 | 4 | 400% |
| KFOR_26 | 13 | 4 | 31% |

## Discussion

The pilot of the Near Real Time Surveillance Tool in the NATO Mission in Kosovo provided important information about the utility of the tool. Key findings were ease of use of the app, the importance of alerts and the need to add syndromic surveillance elements into the tool.

The symptoms reported by the app were consistent with what was expected during the winter season in KFOR with a predominance of respiratory tract presentations such as sore throats and cough. No outbreaks were reported during the pilot so it is not thought that the tool missed anything. However, the tool did aid detection of two incidents; a cluster of skin lesions and two cases of diarrhoea.

It was possible to review the frequency of different combinations of symptoms. This is important as it might be possible to use these combinations to set up algorithms for specific diseases. If, for example, there is a pattern of specific symptoms that is repeated then that might identify a certain disease. In this case the most frequent combination was cough and pain in throat, followed by cough and headache.

The data showed that the most frequent day of reporting was Monday with Sunday being the most infrequent. This was expected as Monday tends to be the busiest day for MTFs but it also showed that reporting took place every day and was not confined to specific days. The number of reports from MTFs followed an expected pattern with the highest number of reports from the MTFs with the largest population at risk.

An assessment of the completeness of the data collected was not possible as the MILMED COE team did not have access to the patient record in each MTF. Comparison with EpiNATO-2 data suggested that there may have been under-reporting by the tool. This suspicion was heightened by the largest number of cases being entered on the day that the MILMED COE team was visiting the MTFs. Further work is required to better understand the completeness of reporting using this tool.

Multiple data entry was a significant issue with clinical staff being asked to enter patient symptoms into the NRTS tool as well as their own patient record. It should be noted that this is routinely required for EpiNATO-2 reporting. The software that runs the tool can receive information from electronic patient records which negates the requirement for manual data entry into the tool however this is not possible on a multinational NATO mission where not all MTFs have electronic patient records. It is, however, possible to combine EpiNATO-2 and NRTS to have one NATO health surveillance tool which would reduce the data entry requirement.

### Key Findings

- Deployment health surveillance is easily collected using an app on a smart device.
- Data analysis requirements are different for different roles within the system. For example: front line clinicians were less interested in the analysis than the JMED team however both wanted to know if there was an outbreak or incident which makes the alert system critical.
- The app was able to detect patterns of presentations that were not immediately apparent to the clinical team. For example: identifying a cluster of patients presenting with papules.
- Most Alerts generated by the system were either for cough or pain in throat. Two cases presenting with these symptoms did not tend to generate any concern as this number was routine for the MTF. Further work is required to better understand the thresholds for triggering Alerts to make them useful.
- Analysis suggested under-reporting in the NRTS app however this was difficult to quantify, and further work is required to better understand compliance with data entry requirements.

### Future Work

There are three areas that need to be tested further:

a. The analysis page needs to be refined following feedback from the KFOR pilot. The new iteration will need to be tested with clinicians.
b. The Alerts need to be refined and thresholds re-set based on the symptoms being seen.
c. There should be only one NATO deployment health surveillance tool – not both near real time and syndromic surveillance (EpiNATO-2). This means combining the two types of surveillance into one tool and piloting the new version.

## Conclusion

The NRTS tool the has potential to deliver the requirement for near real time surveillance set out in the Prague Summit in 2002. Currently, this capability can only be delivered as an addition to patient record systems as many Nations have only paper records. However, with the advent of new electronic patient records, the NRTS tool can be deployed to receive data from the ePR systems negating the need for additional data entry by the MTF. This has been tested on the NATO Coalition Warrior Interoperability Exercises over the last three years and been shown to work.

It is expected that, with further refinements, particularly with the alert system, that the NRTS Tool will be fully deployable and could be integrated with the EpiNATO-2 surveillance tool in the near future. This will provide NATO with near-real time and longer-term information for planning using one tool and gives Commanders increased confidence that the medical information required to detect outbreaks and understand patterns of disease is being exploited.

## Data Availability

All data produced in the present study are available upon reasonable request to the authors

## Notes

### Competing Interest Statement

The authors have declared no competing interest.

### Funding Statement

The study was funded by the NATO Centre of Excellence for Military Medicine

### Author Declarations

The UK Defence Joint Service Publication 536 Pt 2 Annex 1A states that no ethical review is required if the project is not research. The protocol defines this study as an audit and service evaluation which is NOT considered research therefore ethical review was not required in this case.

## References

1. NATO. Prague Summit Declaration [online] 2002. https://www.nato.int/cps/en/natohq/official_texts_19552.htm (accessed 19 July 2023)

2. Caserio-Schönemann C, Meynard JB. Ten years experience of syndromic surveillance for civil and military public health, France, 2004-2014. Euro Surveill. 2015;20(19):pii=21126. https://doi.org/10.2807/1560-7917.ES2015.20.19.21126

3. Meynard, JB., Chaudet, H., Green, A.D. et al. Proposal of a framework for evaluating military surveillance systems for early detection of outbreaks on duty areas. BMC Public Health 8, 146 (2008). https://doi.org/10.1186/1471-2458-8-146

4. Hoysal N, Mccown ME, Fazekas L, Krabbe C. EpiNATO-2: Enhancing Situational Awareness and Overall Force Health Protection While Deployed in the Combined Joint NATO Environment: Describing the Identified 2016 Q Fever Outbreak in Kosovo Force (KFOR). J Spec Oper Med. 2019 Spring;19(1):76–80. doi: 10.55460/EDX8-AQPZ. PMID: 30859532.

5 Holtherm HU. Development of a multinational Deployment Health Surveillance Capability (DHSC) for NATO [online]. 2019. https://military-medicine.com/article/3650-development-of-a-multinational-deployment-health-surveillance-capability-dhsc-for-nato.html (accessed 19 July 2023)

