## Supplemental Information for "Findings from a Pilot of a Near Real Time Disease Surveillance Tool on the NATO Mission in Kosovo"

### Supplementary Information

### Annex A: Symptom List

| Fever and General State Symptoms and Signs | Fever |
| --- | --- |
|  | Excessive Sweating |
|  | Fatigue |
|  | Lymphadenopathy |
| Cardiovascular Symptoms and Signs | Cardiac arrest |
|  | Chest pain |
|  | Palpitations |
| Respiratory and ORL Symptoms and Signs | Respiratory arrest |
|  | Cyanosis |
|  | Difficulty breathing |
|  | Hemoptysis |
|  | Wheezing |
|  | Crepitations |
|  | Cough |
|  | Pain in throat |
|  | Bleeding from nose |
|  | Bronchorrhea |
| Neurological Symptoms and Signs | Disturbance of consciousness |
|  | Disorders of motor control and attention |
|  | Altered sensation |
|  | Headache |
|  | Stiff Neck |
|  | Dizziness |
|  | Photophobia |
|  | Seizure |
|  | Dilirium |
| Ophthalmic Symptoms and Signs | Epiphora |
|  | Conjunctival haemorrhage |
|  | Altered vision |
|  | Conjunctivitis |
|  | Pinpoint pupils |
| Abdominal Symptoms and Signs | Vomiting |
|  | Nausea |
|  | Diarrhea |
|  | Watery diarrhoea |
|  | Abdominal pain |
|  | Hematemesis |
|  | Hematochezia |
|  | Jaundice |
|  | Blood in urine (Hematuria syndrome) |
|  | Anuria |
| Skin Symptoms and Signs | Skin and/or mucous membrane haemorrhage: purpura, bruise, other |
|  | Skin ulcer |
|  | Vesicles |
|  | Papules |
|  | Eruption of skin |
| Psychiatric Symptoms and Signs | Psychotic disorders |
|  | Panic attack |
| Rapid diagnostic test for malaria | Malaria suspected |
| Vital Signs | Temperature |
|  | Blood Pressure |
|  | Normal Pulse Rate |
|  | Respiratory Rate (oberservable entity) |

### Annex B: Evaluation Questions for the NATO Near Real Time Surveillance Tool

The following questions will be discussed with you during the evaluation of the Near Real Time Surveillance Tool on 16-17 Jan 23.

Please DO NOT send us the answers to these questions now but we would be grateful if you could think about your answers before we visit.

**1.0**	**Overview of using the App**

1.1	Please describe how you or your team have used the NRTS app over the last six weeks.

For example:

a.	Have you used it to collect symptoms and signs?

b.	Have you used it for anything else? For example – looking at the summary data for your MTF?

1.2	How easy is the app to use to enter data?

1.3	How easy is it to use the app to access your summary data?

1.4	Can you suggest anything to make data entry into the app easier?

1.5	Can you suggest anything to make the summary data on the app easier to use or understand?

**2.0**	**Impact of the App**

2.1	Did you learn anything from the data you entered into the app? If so what?

For example

a.	Did you discuss the data with the team at the MTF? If so – what did you discuss?

b.	Did you share the data with anyone else? If so – who did you share it with, what did you share, and why did you share it?

2.2	What did you do with the information from the app?

2.3	Were there any actions that resulted from using the data?

**3.0**	**Signs and Symptoms**

3.1	Are any symptoms or signs missing from the list? If so – what?

3.2	Are there any symptoms or signs that could be removed from the list? If so – what?

**4.0**	**EpiNATO-2**

4.1	One of the options is to expand the data being collected on the app to include EpiNATO-2 – what do you think about this?

**5.0**	**Recommendation**

5.1	If you were given the choice, would you continue to use the app even if you did not have to?

Please send any questions to SGM Claudia Biendl

### Annex C: Dummy Patient List

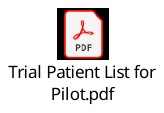

### Annex D: Standard Operating Procedure

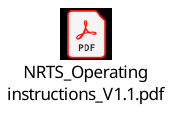
